# Performance of a Computational Phenotyping Algorithm for Sarcoidosis Using Diagnostic Codes in Electronic Medical Records: A Pilot Study from Two Veterans Affairs Medical Centers

**DOI:** 10.1101/2021.02.02.21250980

**Authors:** Mohamed I Seedahmed, Izabella Mogilnicka, Siyang Zeng, Gang Luo, Charles McCulloch, Laura Koth, Mehrdad Arjomandi

**Author notes:** MIS and IM: These authors contributed equally to this work. MA and LK: These authors share senior authorship. **Email Addresses | ORCID iDs: MIS:** IM SZ GL CM LK MA. **Corresponding Author: Mohamed I Seedahmed, MD, MPH**, Division of Pulmonary, Critical care, allergy and Immunology, and Sleep., Department of Medicine, University of California, San Francisco, University of California San Francisco Helen Diller Medical Center, 513 Parnassus Ave., HSE 1314, Box 0111, San Francisco, CA 94143. **Authors’ Contributions:** All authors read and approved the final manuscript. Conceived and designed the study research: MIS, LK, MA Developed study protocol: MIS, LK, MA Worked on the methods: MIS, IM, SZ, CM, LK, MA Analyzed and Interpreted data: MIS, IM, SZ, CM, LK, MA Prepared and/or edit the manuscript: MIS, IM, GL, CM, LK, MA.

## Abstract

**Background:** The accuracy of identifying sarcoidosis cases in electronic medical records (EMR) using diagnostic codes is unknown.

**Methods:** To estimate the statistical performance of using diagnostic codes, ICD-9 and ICD-10 diagnostic codes in identifying sarcoidosis cases in EMR, we searched the San Francisco and Palo Alto Veterans Affairs (VA) medical centers EMR and randomly selected 200 patients coded as sarcoidosis. To further improve diagnostic accuracy, we developed an “index of suspicion” algorithm to identify probable sarcoidosis cases based on clinical and radiographic features. We then determined the positive predictive value (PPV) of diagnosing sarcoidosis by two computational methods using ICD only and ICD plus the “index of suspicion” against the gold standard developed through manual chart review based on the American Thoracic Society (ATS) practice guideline. Finally, we determined healthcare providers’ adherence to the guidelines using a new scoring system.

**Results:** The PPV of identifying sarcoidosis cases in VA EMR using ICD codes only was 71% (95%CI=64.7%-77.3%). The inclusion of our construct of “index of suspicion” along with the ICD codes significantly increased the PPV to 90% (95%CI=85.2%-94.6%). The care of sarcoidosis patients was more likely to be classified as “Fully” or “Substantially” adherent with the ATS practice guideline if their managing provider was a specialist (45% of primary care providers vs. 74% of specialists; P=0.008).

**Conclusions:** Although ICD codes can be used as reasonable classifiers to identify sarcoidosis cases within EMR, using computational algorithms to extract clinical and radiographic information (“index of suspicion”) from unstructured data could significantly improve case identification accuracy.

**Highlights:**

- Identifying sarcoidosis cases using diagnostic codes in EMR has low accuracy.
- “Unstructured data” contain information useful in identifying cases of sarcoidosis.
- Computational algorithms could improve the accuracy and efficiency of case identification in EMR.
- We introduce a new scoring system for assessing healthcare providers’ compliance with the American Thoracic Society (ATS) practice guideline.
- Compliance scoring could help automatically assess sarcoidosis patients’ care delivery.

## INTRODUCTION

Sarcoidosis is a complex disease of unknown etiology that can involve multiple organs, with no universal and standardized measures that fully secure the final diagnosis.[1–3] In fact, only recently the American Thoracic Society (ATS) published its first practice guideline to provide recommendations for diagnosing sarcoidosis and the necessary screening tests.[3] The ATS practice guideline for diagnosis requires the presence of specific clinical and radiographic features, tissue biopsy revealing non-necrotizing granulomas, and exclusion of alternative conditions that can mimic sarcoidosis.[1,3,4]

Electronic medical record (EMR) data are commonly used in research and by healthcare systems, including the Department of Veterans Affairs, to predict outcomes or assess care quality.[5] EMR data are generally captured in two forms: 1) *structured data* such as billing codes like International Classification of Disease (ICD) codes, laboratory test results, procedural codes, and 2) *narrative or unstructured data* such as progress notes, pathology reports, and imaging reports. Unstructured data contain many more details of the clinical conditions, but extracting these details is challenging and time-consuming. In contrast, structured data are easier to search and offer the promise to identify cases computationally using diagnostic codes. But, diagnostic codes can be inaccurate and not easily verifiable, particularly for the case definition of complex diseases such as sarcoidosis.[6–8] Thus, there is a need to develop automated algorithms to verify the final diagnosis of sarcoidosis using unstructured data by incorporating the ATS diagnostic criteria.

The goals of this study are twofold. First, we sought to estimate the statistical performance of using diagnostic codes (ICD-9 and 10) to identify patients with sarcoidosis in the United States Veterans Affairs (VA) EMR through the VA Informatics and Computational Infrastructure (VINCI) via applying a recently published ATS practice guideline. Second, we investigate healthcare providers’ practice patterns and determine how adherent these patterns are with the new practice guideline issued by the ATS. This study will help researchers and healthcare systems understand the healthcare providers’ compliance with these new recommendations and conduct research using administrative databases.

## METHODS

### Study Design

We determined the statistical performance of using diagnostic codes (the International Classification of Diseases, Ninth and Tenth Revisions codes; ICD-9 and ICD-10) to identify patients with sarcoidosis from the EMR. First, by identifying patients with ICD diagnosis code of sarcoidosis, and then performing a comprehensive chart review of the entire unstructured data to ascertain the true diagnosis of sarcoidosis. Furthermore, to improve the diagnostic accuracy using ICD code alone, we generated an “index of suspicion” criteria from unstructured clinical and radiographic data, but not histopathologic data, as a second decision point along with ICD code. We then determined the positive predictive value of both the use of ICD code and ICD code plus “index of suspicion” to determine whether the use of “index of suspicion” could improve the accuracy of identifying sarcoidosis cases in EMR regardless of tissue biopsy availability. Finally, we investigated the relevant healthcare utilization among the identified patients to determine healthcare providers’ adherence to the ATS practice guideline.

### Data Source

This is a retrospective cohort study of electronic medical records (EMR) available from the Veterans Affairs Informatics and Computational Infrastructure (VINCI). Developed by the VA Health Services Research & Development (HSR&D) to improve veterans’ healthcare, VINCI provides access to comprehensive and integrated veterans’ national datasets that are de-identified and the necessary computational and analytical tools in a secure, high-performance computing environment [9]. The University of California San Francisco Institutional Review Board and the Veterans Health Administration Research and Development Committee approved this study.

### Data Collection

We searched the EMR data in VINCI from 1989 to 2019 and identified all patients coded as sarcoidosis cases in the VA healthcare system, as defined by documentation of the ICD-9 and 10 codes of 135 and D.86 (including all subcodes), respectively. Data were extracted through executing Structured Query Language (SQL) queries in a SQL Server 2017 database.

We then performed a comprehensive and thorough manual chart review of the unstructured data (available clinical notes, pathology reports, radiology reports, and laboratory test results) to determine the accuracy of the ICD-9 or 10 code-based diagnosis of sarcoidosis among cases. To limit the required chart review to a manageable level, we reviewed a total of 200 cases out of the 14,833 identified. Because our access to the detailed medical records was limited to two VA medical centers (San Francisco VA [SFVA] and Palo Alto Medical Centers [PAVA]), the 200 patients coded as sarcoidosis were selected from those two centers. We stratified the list of sarcoidosis cases from the two centers by site and used the “lottery” method to select 100 patients from each site randomly.[10]

Two independent reviewers (MIS, IM) confirmed the diagnoses of sarcoidosis by performing a manual chart review of the 200 cases in the VA EMR, Computerized Patient Record System (CPRS), and based on the ATS diagnostic criteria (clinical, radiographic, pathological findings, and exclusion of other causes).[3] This approach was considered the “gold standard” methodology for diagnostic accuracy.

Given that in clinical practice, not all suspected patients will have a tissue biopsy, we generated an “index of suspicion” for sarcoidosis to identify patients with probable sarcoidosis based on clinical and radiographic information and regardless of the availability of biopsy data. The “index of suspicion” was defined based on the interpretation of available unstructured data from both clinical and radiographic findings without the inclusion of tissue biopsy results if available and was used to determine the likelihood of an individual having sarcoidosis. Upon completing the chart review, each patient was assigned to one of two groups of “high index of suspicion” if the clinical and radiological presentations were supportive of sarcoidosis, or “low index of suspicion” if clinical and radiological findings were not consistent with sarcoidosis. We then classified the patients into three groups: (1) Patients with a high index of suspicion and documented histopathological evidence of non-necrotizing granulomas were categorized as “probable sarcoidosis with confirmed biopsy”; (2) Those with a high index of suspicion and either no documented biopsy in the EMR or a biopsy showing no histopathological evidence of non-necrotizing granulomas were categorized as “probable sarcoidosis without confirmed biopsy”; and (3) those with a low index of suspicion were classified as “unlikely sarcoidosis.”

This approach was used to compare the statistical performance of the two methods (ICD code alone versus ICD code with “index of suspicion”) in identifying sarcoidosis patients from the EMR. As we started with a random sample of those with sarcoidosis diagnostic codes (ICD codes 135 or D86), the further restriction of the sample to those with “high index of suspicion” is still a random sample of the combination of both ICD codes and “index of suspicion.”

### Disease-related Variables

Organ involvement was assessed based on the clinical history obtained from physicians’ notes and imaging and biopsy reports available in CPRS. For this assessment, to adjust for the variability in providers’ documentation, we adapted a set of criteria previously introduced in the NIH-sponsored Genomic Research in Alpha-1 Antitrypsin Deficiency and Sarcoidosis (GRADS) Study.[11]

We collected the following data from chart review: clinical site (SFVA or PAVA), gender, race, ICD-9 and 10 code for sarcoidosis (135 and D86), index date of ICD code (entry date of the ICD code), specialty visits (specialty notes available), the pathological diagnosis from any available biopsy, organ involvement (as described in **Figure 2**), Scadding staging of chest x-ray (as described in radiology reports), history of bilateral hilar lymphadenopathy (based on radiology reports and clinical notes), pulmonary function test (**PFT**) pattern (as reported in PFT reports), vitamin D levels (25 [OH]D and 1,25[OH]D), and finally the treatment status of sarcoidosis. Besides, we checked whether other imaging and laboratory reports were performed and available in EMR as part of diagnostic workup and sarcoidosis management; including echocardiography, 12-lead electrocardiogram (**ECG**), chest x-ray, chest computed tomography (**CT**), cardiac magnetic resonance imaging (**MRI**) or positron emission tomography (**PET**)/ CT, or abdominal CT scan or ultrasound.

Pathological diagnoses were categorized into “primary” histopathological if it was available in the pathology report domains, or “secondary” if the data was only available in clinical note domains due to either a remote history of biopsy or the biopsy being performed outside of the VA. The PFT reports at SFVA and PAVA used Crapo reference equations to calculate the Lower Limit of Normal (**LLN**) values for spirometry and lung volume measurements.

Using the clinical data from chart abstraction, we classified the patients into the clinical phenotypes proposed by the GRADS study with some modifications.[11] Because of the variability of available data needed to confirm multiple organ involvement from EMR chart review, we considered three or more organ involvement (instead of five or more as defined in the GRADS study) to categorize “multiorgan phenotype.”

Finally, we created a scoring system to estimate the physicians’ compliance with the recently published ATS clinical practice guideline.[3] For this analysis, four variables were used in the scoring system: (1) having an ophthalmology visit, (2) 12-lead EKG, (3) chest X-ray, and (4) vitamin D measurements (25-(OH) D and 1,25(OH) D) within one year after the index date of ICD code. Each variable was given a score of three, and each patient could have a total score of 12. We classified the physicians’ compliance into four categories based on the score: “Fully,” “Substantially,” “Partially,” or “Non-compliant” for scores of 12, 9-11, 5-8, or <5 points, respectively. This scoring system was applied to all probable sarcoidosis patients with or without histopathology confirmation. Furthermore, the role of primary versus specialty care evaluation and management was examined by comparing the healthcare providers’ compliance score.

### Statistical Analyses

All statistical analyses were performed with R software (RStudio, version 1.2.5, Foundation for Statistical Computing, Vienna, Austria). Descriptive statistics were computed to summarize the data. Categorical variables were presented as the frequency in percentages and continuous data as means and standard deviations. We estimated the positive predictive value (PPV) of the two computational diagnostic criteria for sarcoidosis described above (ICD codes only and ICD codes along with “index of suspicion”). The PPV of the criterion of using ICD code only was calculated as the number of patients verified to have sarcoidosis by chart review (“gold standard” or true positives) divided by the total number of patients with an ICD-9 or 10 diagnostic code of sarcoidosis. The PPV of the criterion of using ICD codes and “index of suspicion” use was calculated as the number of patients verified to have sarcoidosis (“gold standard” or true positives) divided by the total number of patients with a high index of suspicion. We computed the 95% confidence intervals (CI) using the exact binomial method. For our estimates, the significance was defined as P < 0.05.

To determine whether it is worth interpreting the developed contingency table that compares the healthcare providers’ compliance score by primary versus specialty care, we computed the chi-squared test of significance.

## RESULTS

### Patients’ Characteristics

A total of 14,833 patients with at least one ICD-9 or 10 diagnostic code of sarcoidosis were identified (**Figure 1**). The study cohort included 200 patients identified by ICD codes of sarcoidosis: 169 with ICD-9 code of 135 and 39 with ICD-10 code of D.86 (**Table 1**). Of the 200 patients, 158 had a “high index of suspicion” for sarcoidosis based on clinical and radiographic findings. Of those, 142 had confirmed sarcoidosis based on histopathological evidence of non-necrotizing granuloma and were classified as “probable sarcoidosis with confirmed biopsy.” The remaining 16 patients with a “high index of suspicion” did not undergo a biopsy and were classified as “probable sarcoidosis without confirmed biopsy” (**Figure 2**). No patient had nondiagnostic biopsy results for sarcoidosis.

**Table 1.**
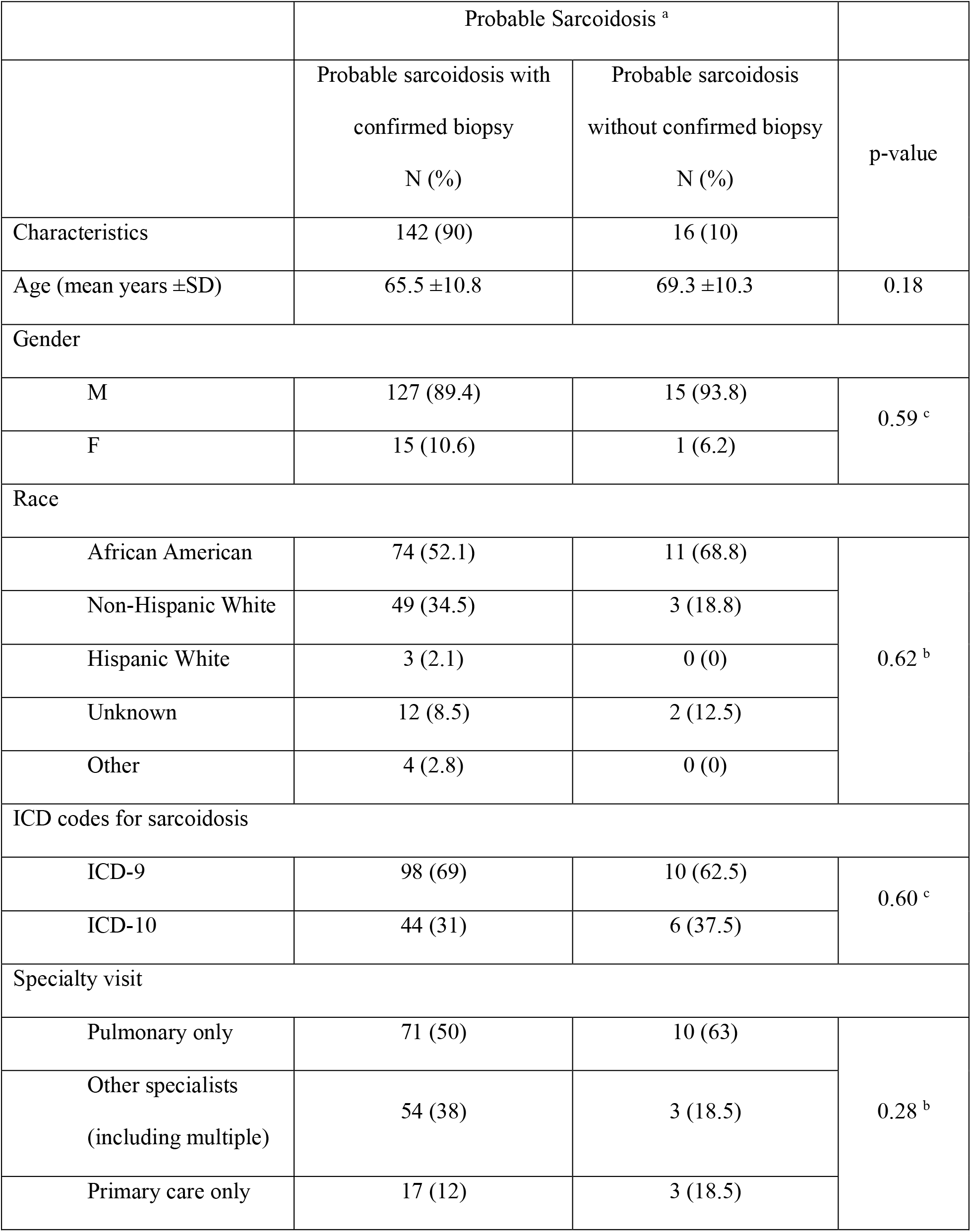
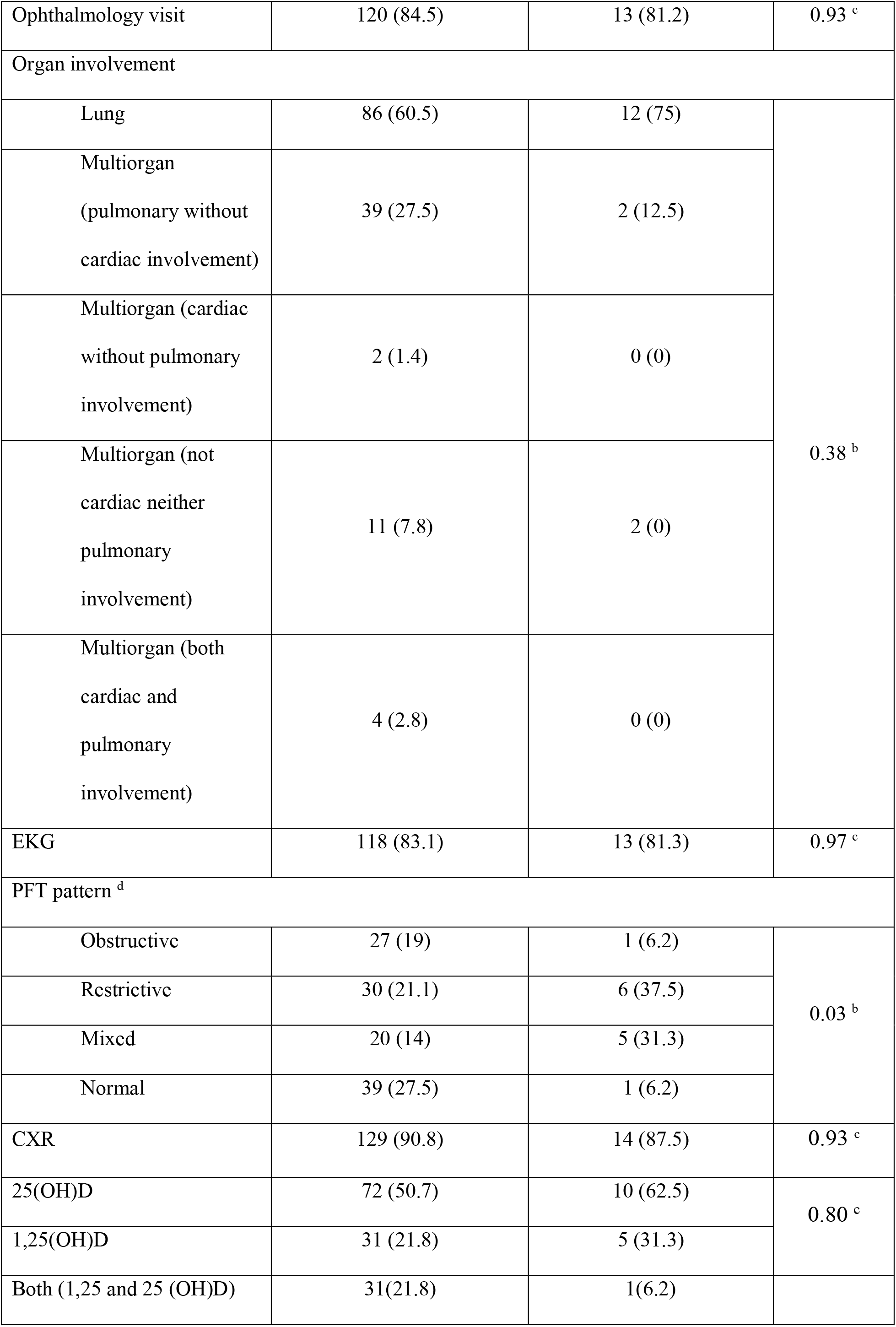
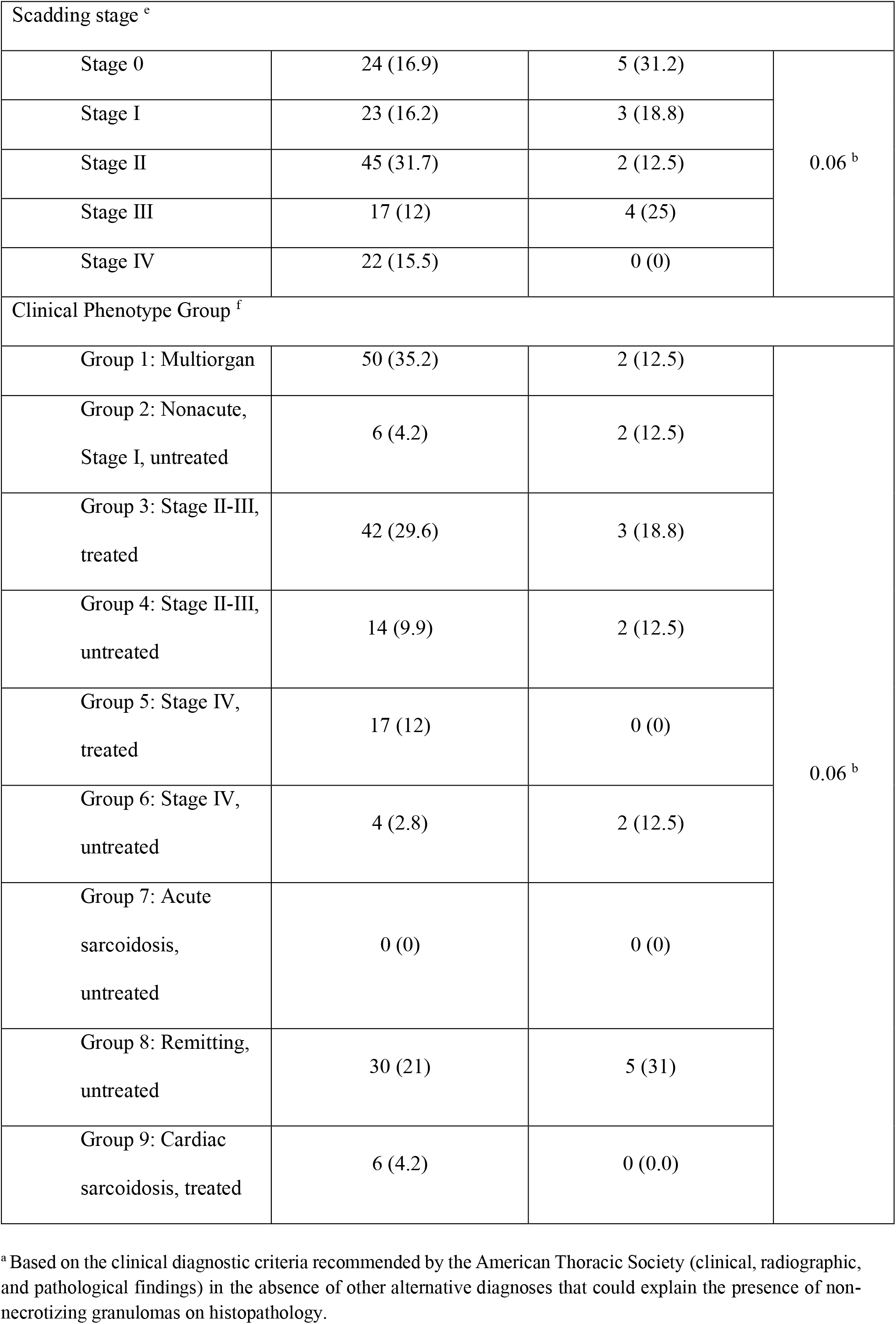
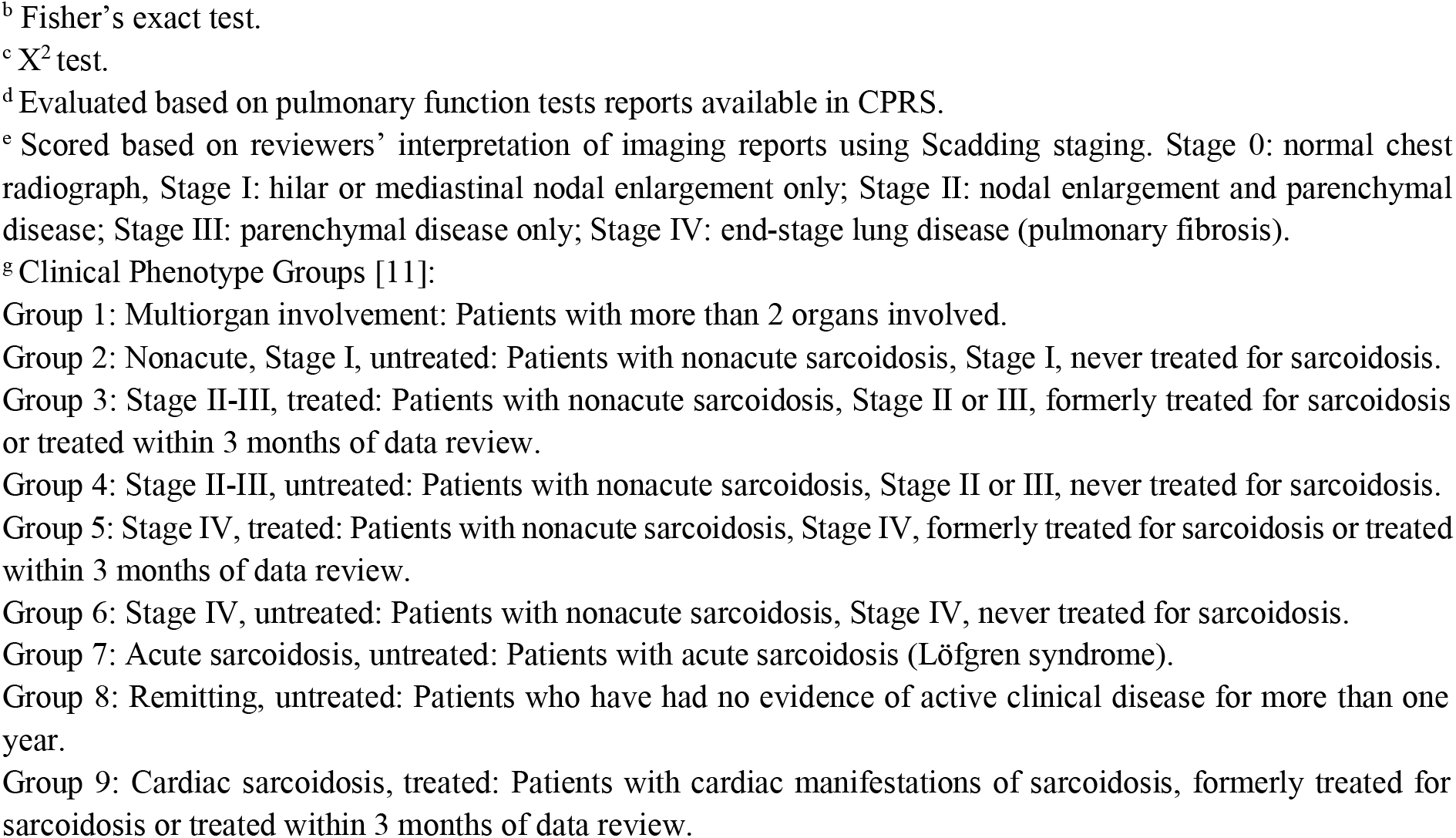
Distribution of characteristics and clinical phenotype groups of patients with probable sarcoidosis ^a^ (with and without confirmed biopsy)

|  | Probable Sarcoidosis <sup>a</sup> |  |  |
| --- | --- | --- | --- |
|  | Probable sarcoidosis with<br>confirmed biopsy<br>N (%) | Probable sarcoidosis<br>without confirmed biopsy<br>N (%) | p-value |
| Characteristics | 142 (90) | 16 (10) |  |
| Age (mean years ±SD) | 65.5 ±10.8 | 69.3 ±10.3 | 0.18 |
| Gender |  |  |  |
| M | 127 (89.4) | 15 (93.8) | 0.59 <sup>c</sup> |
| F | 15 (10.6) | 1 (6.2) |  |
| Race |  |  |  |
| African American | 74 (52.1) | 11 (68.8) | 0.62 <sup>b</sup> |
| Non-Hispanic White | 49 (34.5) | 3 (18.8) |  |
| Hispanic White | 3 (2.1) | 0 (0) |  |
| Unknown | 12 (8.5) | 2 (12.5) |  |
| Other | 4 (2.8) | 0 (0) |  |
| ICD codes for sarcoidosis |  |  |  |
| ICD-9 | 98 (69) | 10 (62.5) | 0.60 <sup>c</sup> |
| ICD-10 | 44 (31) | 6 (37.5) |  |
| Specialty visit |  |  |  |
| Pulmonary only | 71 (50) | 10 (63) | 0.28 <sup>b</sup> |
| Other specialists<br>(including multiple) | 54 (38) | 3 (18.5) |  |
| Primary care only | 17 (12) | 3 (18.5) |  |
| Ophthalmology visit | 120 (84.5) | 13 (81.2) | 0.93 <sup>c</sup> |
| Organ involvement |  |  |  |
| Lung | 86 (60.5) | 12 (75) | 0.38 <sup>b</sup> |
| Multiorgan<br>(pulmonary without<br>cardiac involvement) | 39 (27.5) | 2 (12.5) |  |
| Multiorgan (cardiac<br>without pulmonary<br>involvement) | 2 (1.4) | 0 (0) |  |
| Multiorgan (not<br>cardiac neither<br>pulmonary<br>involvement) | 11 (7.8) | 2 (0) |  |
| Multiorgan (both<br>cardiac and<br>pulmonary<br>involvement) | 4 (2.8) | 0 (0) |  |
| EKG | 118 (83.1) | 13 (81.3) | 0.97 <sup>c</sup> |
| PFT pattern <sup>d</sup> |  |  |  |
| Obstructive | 27 (19) | 1 (6.2) | 0.03 <sup>b</sup> |
| Restrictive | 30 (21.1) | 6 (37.5) |  |
| Mixed | 20 (14) | 5 (31.3) |  |
| Normal | 39 (27.5) | 1 (6.2) |  |
| CXR | 129 (90.8) | 14 (87.5) | 0.93 <sup>c</sup> |
| 25(OH)D | 72 (50.7) | 10 (62.5) | 0.80 <sup>c</sup> |
| 1,25(OH)D | 31 (21.8) | 5 (31.3) |  |
| Both (1,25 and 25 (OH)D) | 31(21.8) | 1(6.2) |  |

| Scadding stage <sup>e</sup> |  |  |  |
| --- | --- | --- | --- |
| Stage 0 | 24 (16.9) | 5 (31.2) | 0.06 <sup>b</sup> |
| Stage I | 23 (16.2) | 3 (18.8) |  |
| Stage II | 45 (31.7) | 2 (12.5) |  |
| Stage III | 17 (12) | 4 (25) |  |
| Stage IV | 22 (15.5) | 0 (0) |  |
| Clinical Phenotype Group <sup>f</sup> |  |  |  |
| Group 1: Multiorgan | 50 (35.2) | 2 (12.5) | 0.06 <sup>b</sup> |
| Group 2: Nonacute, Stage I, untreated | 6 (4.2) | 2 (12.5) |  |
| Group 3: Stage II-III, treated | 42 (29.6) | 3 (18.8) |  |
| Group 4: Stage II-III, untreated | 14 (9.9) | 2 (12.5) |  |
| Group 5: Stage IV, treated | 17 (12) | 0 (0) |  |
| Group 6: Stage IV, untreated | 4 (2.8) | 2 (12.5) |  |
| Group 7: Acute sarcoidosis, untreated | 0 (0) | 0 (0) |  |
| Group 8: Remitting, untreated | 30 (21) | 5 (31) |  |
| Group 9: Cardiac sarcoidosis, treated | 6 (4.2) | 0 (0.0) |  |

<sup>a</sup> Based on the clinical diagnostic criteria recommended by the American Thoracic Society (clinical, radiographic, and pathological findings) in the absence of other alternative diagnoses that could explain the presence of non-necrotizing granulomas on histopathology.
<sup>b</sup> Fisher's exact test.
<sup>c</sup> X<sup>2</sup> test.
<sup>d</sup> Evaluated based on pulmonary function tests reports available in CPRS.
<sup>e</sup> Scored based on reviewers' interpretation of imaging reports using Scadding staging. Stage 0: normal chest radiograph, Stage I: hilar or mediastinal nodal enlargement only; Stage II: nodal enlargement and parenchymal disease; Stage III: parenchymal disease only; Stage IV: end-stage lung disease (pulmonary fibrosis).
<sup>g</sup> Clinical Phenotype Groups [11]:
Group 1: Multiorgan involvement: Patients with more than 2 organs involved.
Group 2: Nonacute, Stage I, untreated: Patients with nonacute sarcoidosis, Stage I, never treated for sarcoidosis.
Group 3: Stage II-III, treated: Patients with nonacute sarcoidosis, Stage II or III, formerly treated for sarcoidosis or treated within 3 months of data review.
Group 4: Stage II-III, untreated: Patients with nonacute sarcoidosis, Stage II or III, never treated for sarcoidosis.
Group 5: Stage IV, treated: Patients with nonacute sarcoidosis, Stage IV, formerly treated for sarcoidosis or treated within 3 months of data review.
Group 6: Stage IV, untreated: Patients with nonacute sarcoidosis, Stage IV, never treated for sarcoidosis.
Group 7: Acute sarcoidosis, untreated: Patients with acute sarcoidosis (Löfgren syndrome).
Group 8: Remitting, untreated: Patients who have had no evidence of active clinical disease for more than one year.

**Figure 1.**
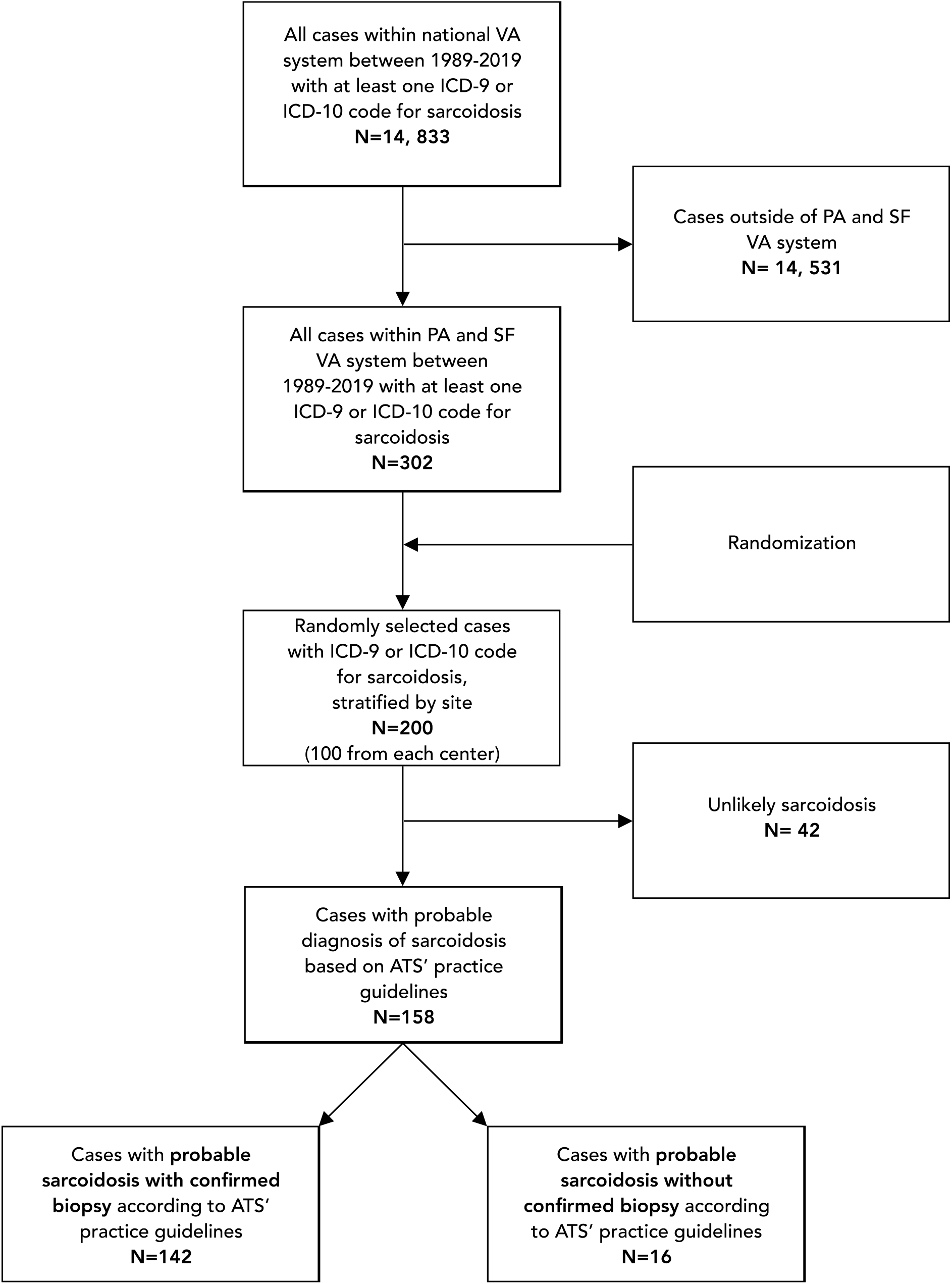
STROBE flow chart. Selection criteria for probable sarcoidosis cases. <u>Abbreviations:</u> ATS, American Thoracic Society; ICD, International Classification of Diseases; PA, Palo Alto; SF, San Francisco; VA, Veterans Affairs

**Figure 2.**
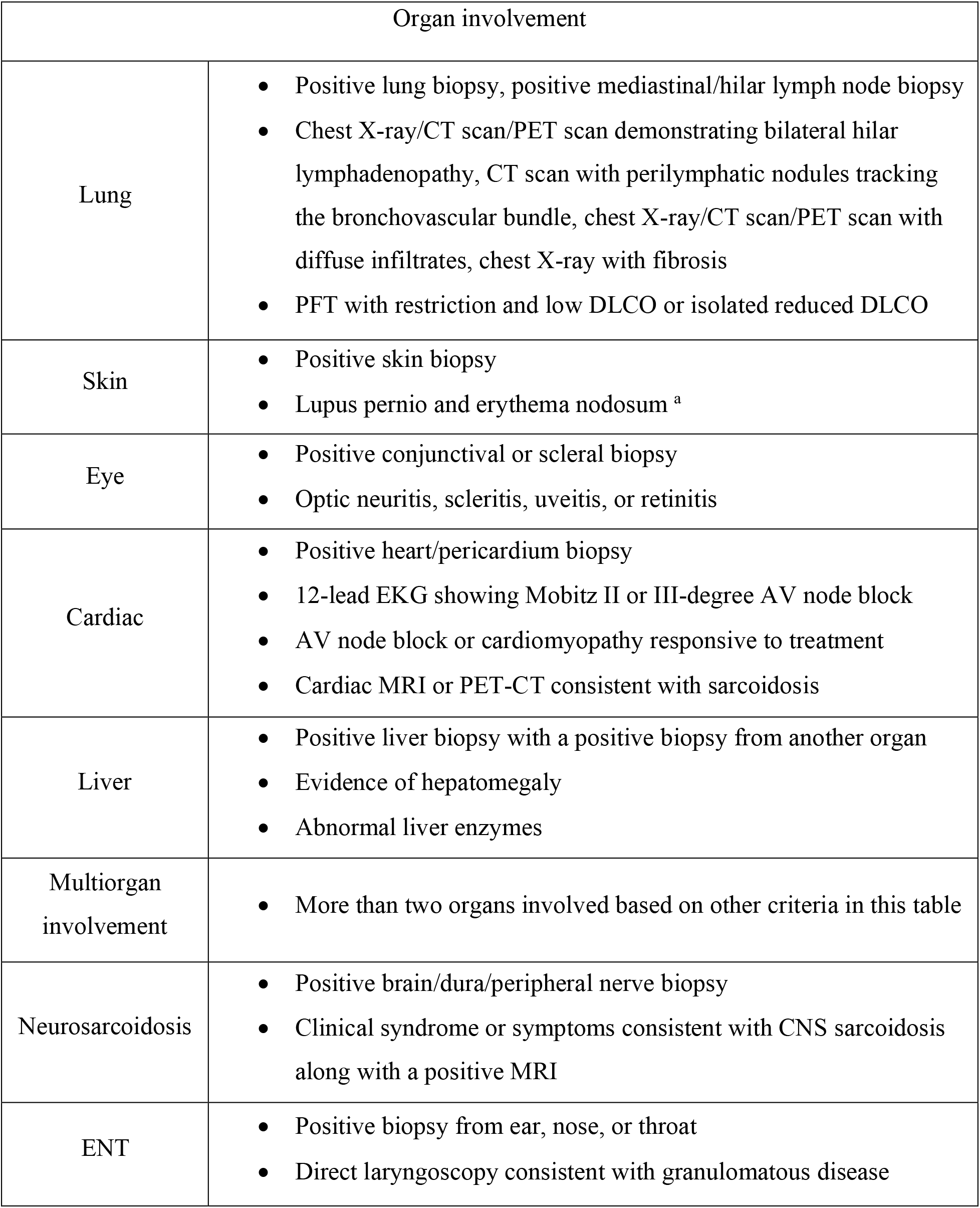
Organ involvement assessment for probable sarcoidosis with and without confirmed biopsy ^a^. <u>Abbreviations:</u> AV, atrioventricular; CNS, central nervous system; CT, computed tomography; CXR, chest radiograph; DLCO, diffusing capacity for carbon monoxide; EKG, electrocardiogram; ENT, ears, nose and throat; MRI, magnetic resonance imaging; PET, positron emission tomography; PFT, pulmonary function test. ^a^ No biopsy is needed for acute skin sarcoidosis.

Among those patients with probable sarcoidosis (with and without confirmed biopsy), 90% (142/158) were males (**Table 1**), and there was a higher representation of African Americans than non-Hispanic Whites (54% (85/158) vs. 33% (52/158), respectively). Overall, 90% (143/158) had a predominant pulmonary phenotype, and 29% (45/158) had a multi-organ disease that included pulmonary. Among those with pulmonary phenotype, 28% (36/129), 22% (28/129), and 19% (25/129) had restrictive, obstructive, and mixed lung function patterns, respectively. The majority were in Scadding stage II (32%, 47/145), followed by Stage 0 and Stage 1 (20% (29/145) and 18% (26/145), respectively). There was no significant difference in age between those who did and those who did not have a biopsy performed to diagnose sarcoidosis (mean age= 65.5 vs. 69.3, P= 0.18, respectively). In terms of clinical phenotypes, the most common phenotype was multiorgan (33%, 52/158), followed by stage II or III treated (29%, 45/158). Our study cohort did not include any individuals with acute presentation (acute, untreated). Some patients with remitting phenotype overlapped with Groups 2 and 4 (**Table 1**) (**Figure 3**).

**Figure 3.**
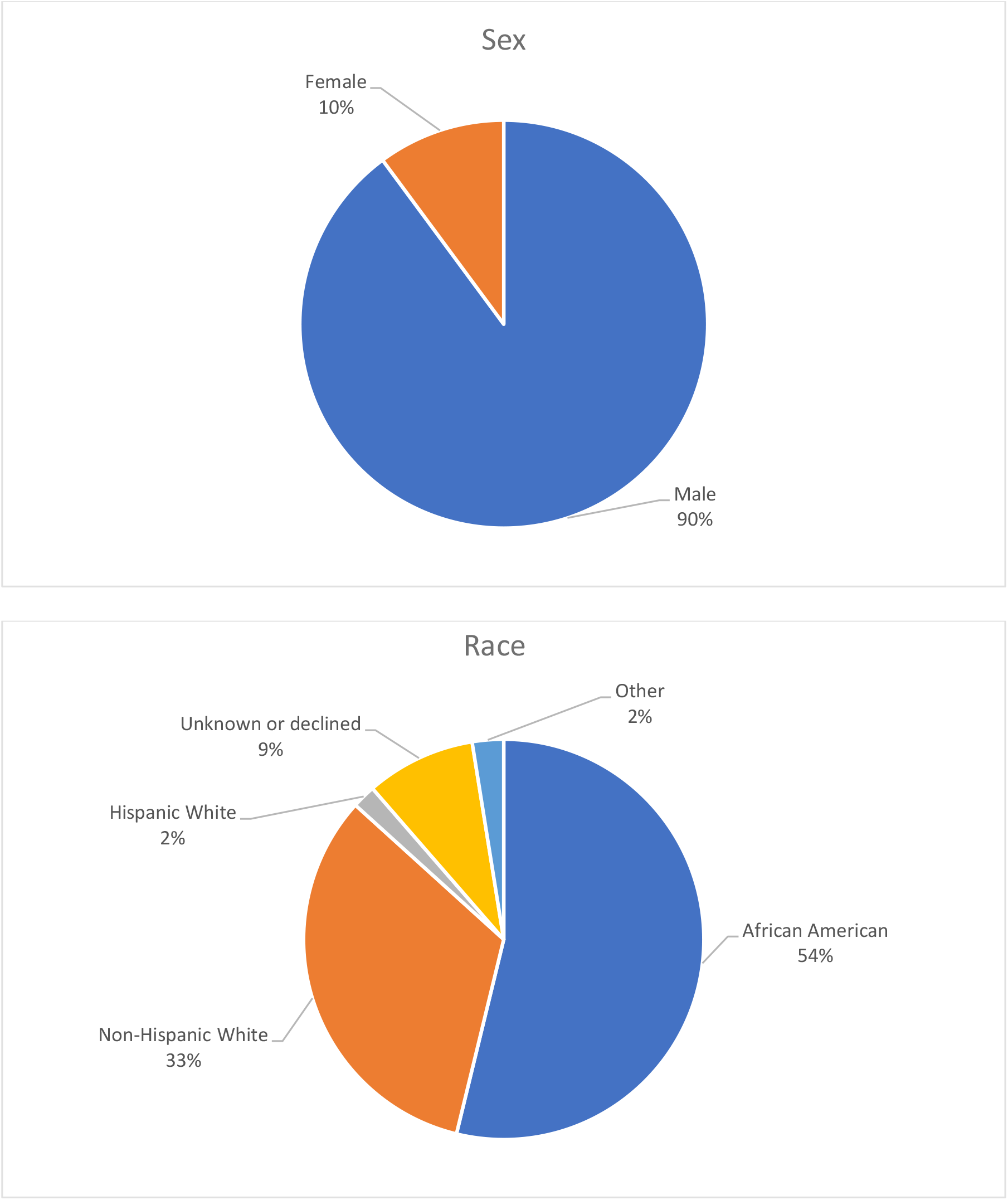
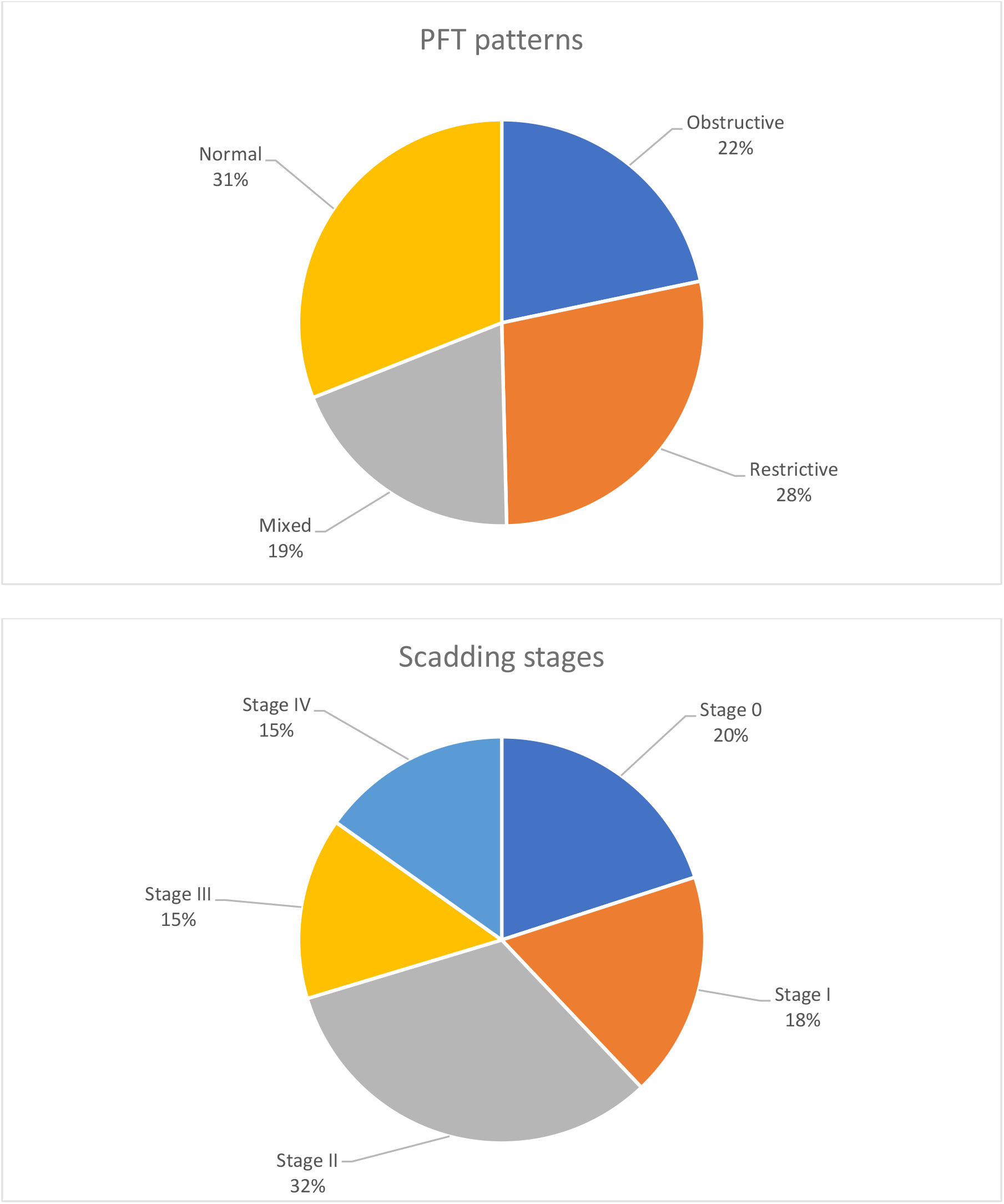
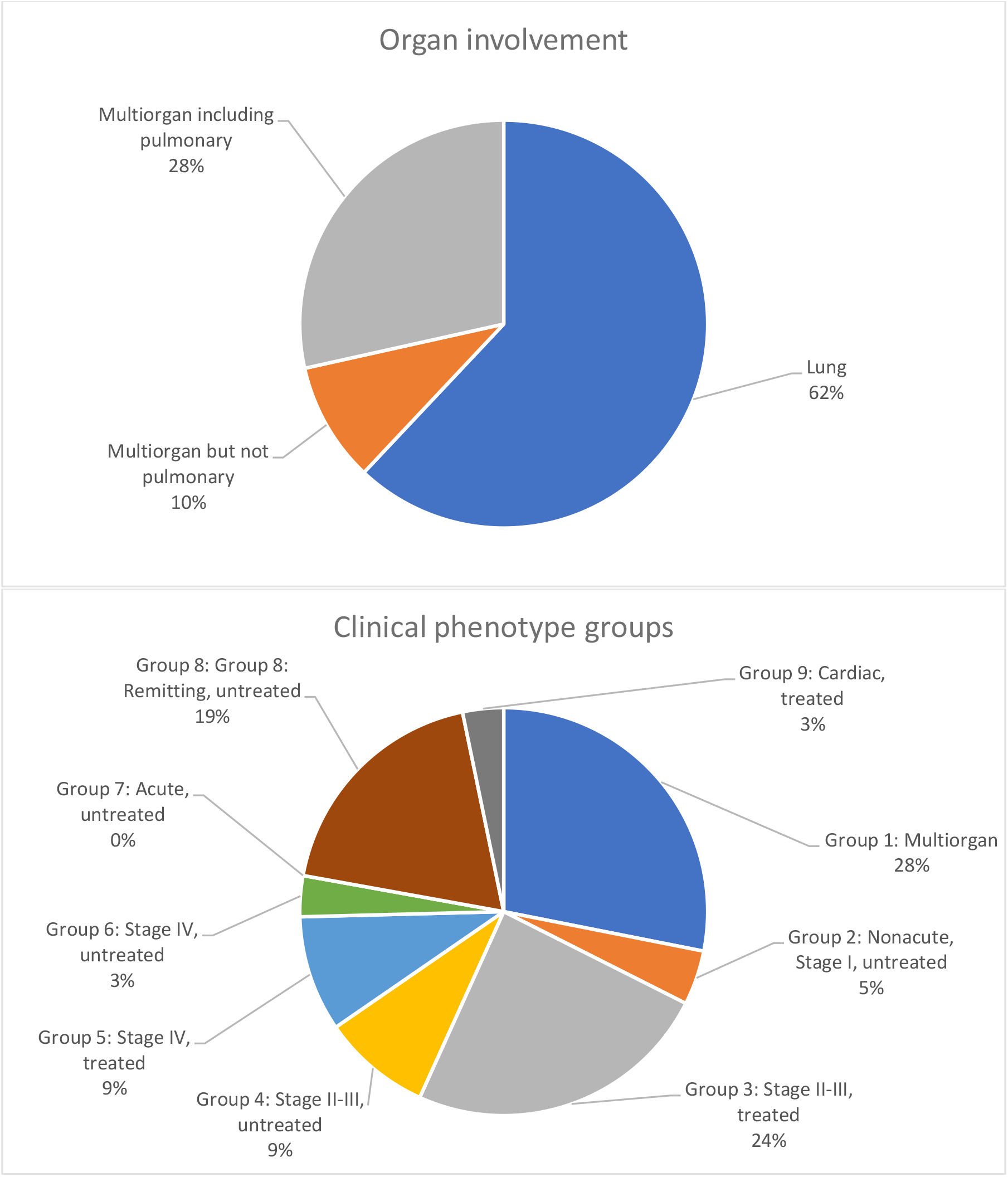
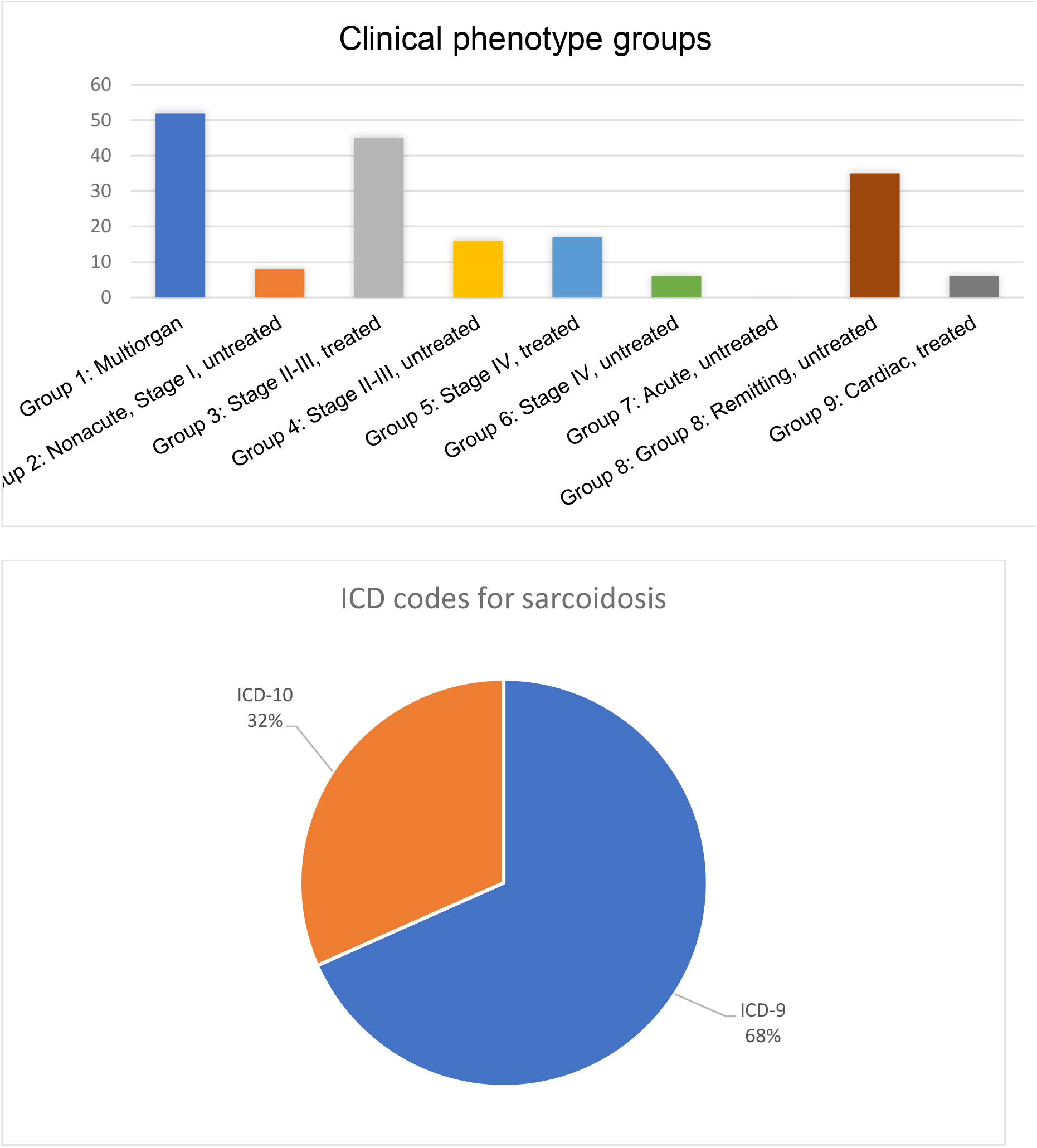
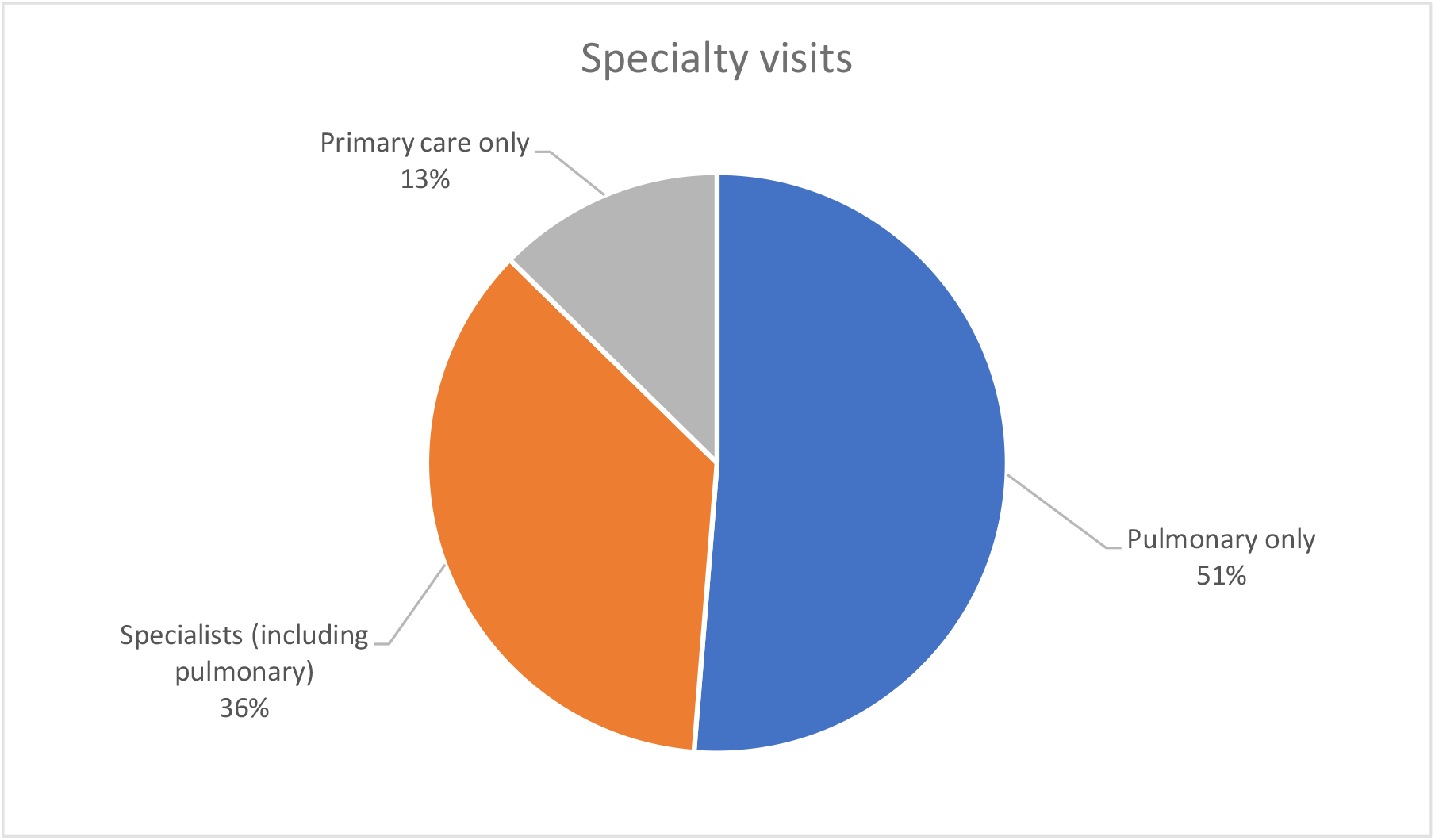
Distribution of characteristics among patients with probable sarcoidosis with and without confirmed biopsy. <u>Abbreviations:</u> PFT, pulmonary function test; ICD, International Classification of Diseases

### Diagnostic Accuracy of ICD Codes

We then calculated the positive predictive value (PPV) of using ICD codes to identify VA patients that met the ATS definition of sarcoidosis from the VINCI database. For this calculation, we used the curated dataset of 200 patients. The estimated PPV of using only ICD codes was 71% (95%CI= 64.7%-77.3%). The inclusion of our construct of “index of suspicion” along with the ICD codes into the identification of sarcoidosis increased the PPV estimate significantly to 90% (95%CI= 85.2%-94.6%) (**Table 2**), as the initial sarcoidosis cohort was restricted to the patients with “high index of suspicion.”

**Table 2.** Contingency 2×2 table of using ICD and index of suspicion for sarcoidosis cases identification.

| Patients with positive ICD 9 or ICD-10 for sarcoidosis | Index of Suspicion <sup>a</sup> | Sarcoidosis confirmed with a biopsy |  |  |
| --- | --- | --- | --- | --- |
|  |  | Yes <sup>b</sup> | No <sup>c</sup> | Total |
|  | High index of suspicion | 142 <sup>d</sup> | 16 <sup>e</sup> | 158 |
|  | Low index of suspicion | 0 <sup>f</sup> | 42 <sup>g</sup> | 42 |
|  | Total | 142 | 58 | 200 |

<sup>a</sup> Index of suspicion for sarcoidosis based on both clinical and radiographic evidence but not biopsy.
<sup>b</sup> Available biopsies with primary or secondary histopathological reports.
<sup>c</sup> No biopsies were ordered or available in the electronic medical record.
<sup>d</sup> Probable sarcoidosis group with histopathological evidence of non-necrotizing granuloma (NNG) = true positives
<sup>e</sup> Probable sarcoidosis group without histopathological evidence of NNG= false positives
<sup>f</sup> No sarcoidosis group due to lack of sufficient clinical and radiological features consistent with sarcoidosis even in the presence of the histopathological evidence of NNG = false negatives
<sup>g</sup> No sarcoidosis group due to lack of sufficient clinical and radiological features consistent with sarcoidosis, in addition to the absence of the histopathological evidence of NNG= true negatives

### Providers’ Compliance with the ATS Clinical Practice Guideline

Among those with probable sarcoidosis (with and without biopsy), 13% were managed by primary care providers only, 51% (81/158) were managed by pulmonary physicians only, and pulmonary physicians managed 36% (57/158) along with other specialties such as ophthalmology or cardiology. The specialty care visits occurred in the context of diagnosis or management of the disease.

Within one year of the entry date of the ICD code of sarcoidosis, 91% (143/158) of patients had at least one chest x-ray, 83% (131/158) had at least one EKG, and 84% (133/158) had at least one visit to an ophthalmologist. In addition, 53% (82/158) had 25(OH)D, and 23% (36/158) had 1, 25(OH)D, including 20% (32/158) who had both measurements (**Table 1**).

Among the patients with probable sarcoidosis (with and without biopsy), 70% (111/158) had managing providers who were “Fully” or “Substantially” compliant with the ATS practice guideline based on our scoring system (**Table 3**). The majority of those patients, 92% (102/111), were managed by specialists, including pulmonary physicians, while primary care providers managed 8% (9/111). Among the 30% (47/158) patients whose care only met the “Partially compliant” or “Non-compliant” definition based on our scoring system, 23% (11/47) were managed by primary care providers (**Figure 4**). This result indicates that the care of sarcoidosis patients was more likely to be classified as “Fully” or “Substantially” adherent with the ATS practice guideline if their managing provider was a specialist (45% [9/20] of primary care providers vs. 74% [102/138] of specialists, P= .008) (**Table 4**).

**Table 3.** Healthcare providers’ compliance with the ATS clinical practice guideline.

| Healthcare providers' compliance with<br>ATS practice guidelines <sup>a</sup><br>N (%) | Probable sarcoidosis<br>with confirmed biopsy<br>N=142 | Probable sarcoidosis<br>without confirmed biopsy<br>N= 16 |
| --- | --- | --- |
| Fully compliant | 26 (18.3) | 1 (6.25) |
| Substantially compliant | 76 (53.5) | 8 (50) |
| Partially compliant | 31 (21.9) | 6 (37.5) |
| Non-compliant | 9 (6.3) | 1 (6.25) |

<sup>a</sup> Developed with modifications based on recently published ATS practice guideline. The scoring system has been developed based on the availability of the following within one year of the diagnosis: ophthalmology visit = 3 points, 12-leads EKG = 3 points, CXR = 3 points, vitamin D (25-OH + 1,25-OH = 3 points; 25(OH)D or 1,25(OH)D = 2 points, otherwise zero points). Each variable weight a score of 3, with a total score of 12. Each patient was given a final score, and the purpose was to classify the degree of the providers' compliance into four categories based on the score:
- o *Fully compliant*: if the score = 12 - o *Substantially compliant*: if the score = 9-11 - o *Partially compliant*: if the score = 5-8 - o *Non-compliant*: if the score <5.

**Table 4.** Contingency 2×2 table of the healthcare providers’ compliance score by primary versus specialty care ^a^.

| Healthcare providers' compliance with<br>ATS practice guideline scored by our<br>classification | Healthcare providers |  |  |
| --- | --- | --- | --- |
|  | Specialists | Primary care<br>physicians | Total |
| "Fully" or "Substantially"<br>compliant | 102 | 9 | 111 |
| "Partially" or "Non" compliant | 36 | 11 | 47 |
| Total | 138 | 20 | 158 |

<sup>a</sup> $\chi^2$ statistic= 6.9877, P-value= 0.008.

**Figure 4.**
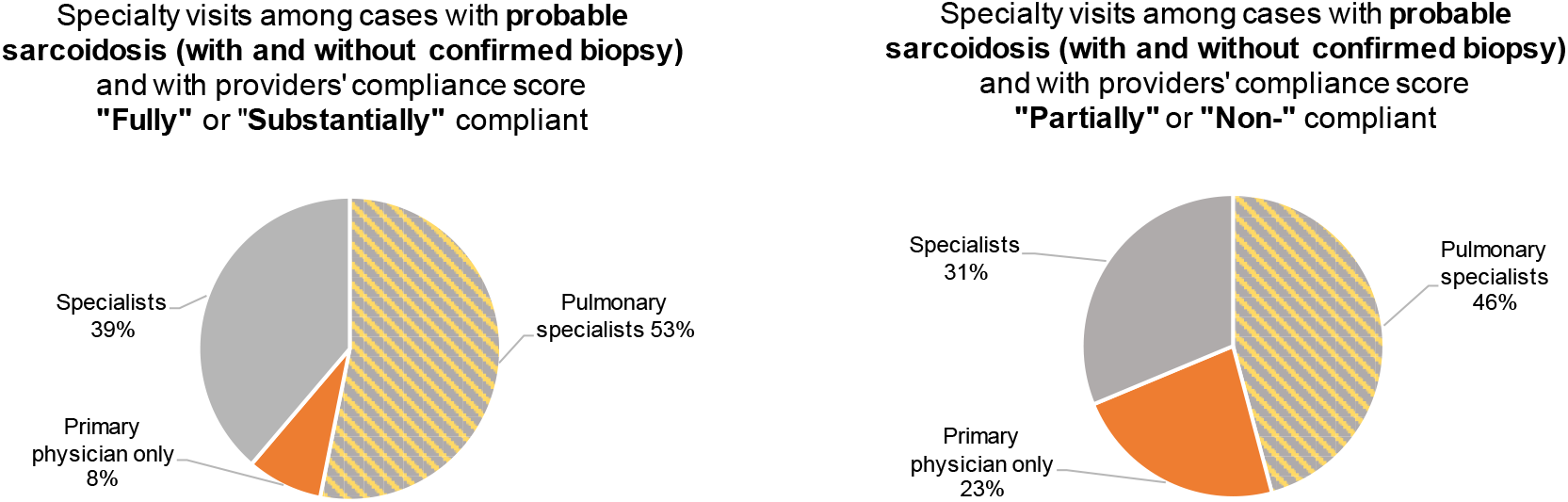
Comparison of specialty visits across cases with probable sarcoidosis (with and without confirmed biopsy) and with healthcare providers classified as “Fully” or “Substantially” compliant vs. “Partially” or “Non-” compliant using our compliance scoring system.

## DISCUSSION

In this retrospective cohort study of Department of Veterans Affairs EMR, we evaluated 200 randomly selected patients diagnosed with sarcoidosis from the San Francisco and Palo Alto Veterans Affairs Medical Centers. We found that using ICD-9 and 10 codes to identify sarcoidosis patients within EMR has a relatively low accuracy (PPV of 71% [95%CI= 64.7%-77.3%]) of detecting patients with “probable sarcoidosis” as defined by the ATS clinical practice guideline. We also demonstrated that including an “index of suspicion” that incorporates only the clinical and radiographical data allows for a significant increase in diagnostic accuracy (PPV of 90% [85.2%-94.6%]), as the initial sarcoidosis cohort was restricted to the patients with “high index of suspicion.” This “index of suspicion,” which was developed based on the ATS clinical practice guideline [3], could be executed by manual chart review of the available clinical and radiographic data and could potentially be adapted for automated chart review algorithms using natural language processing and machine learning in EMR. Furthermore, we found that the care provided by specialists was more likely to be “Fully” or “Substantially” compliant with the ATS practice guideline compared to primary care providers.

This randomly selected cohort of 200 Veterans with sarcoidosis diagnosis consisted of 90% men and 10% women. While the sex distribution in our study was different from the ACCESS study[12], it is closely reflective of the veterans’ population demographics.[13] This study further confirmed the higher prevalence of sarcoidosis in African-Americans (54%) compared to Caucasians (33%), which has been previously reported by many other sarcoidosis epidemiology studies [14–18]. At the same time, the study population was racially diverse, highlighting the potential utility of the VA EMR for studying sarcoidosis in populations of people of color.[19]

The novelty of this study is the development of a scoring system to objectively measure physicians’ compliance with the latest ATS practice guideline. Adherence to recommendations might allow for earlier interventions in sarcoidosis management and ultimately improve outcomes by delaying or preventing the development of morbid phenotypes. Variables used in this scoring system included ophthalmology visit, 12-lead EKG, chest x-ray, and vitamin D levels measurements within one year of the entry date of the ICD code. These variables were chosen based on their utility and importance.[3] Not all tests and measurements discussed in the guidelines have been included in our scoring system. Yet, we believe it can be easily modified to include other clinical variables. Notably, a longitudinal analysis of the association between physicians’ compliance with the recommendations and clinical outcomes in sarcoidosis could provide direction for future investigations and ultimately help better guide clinical practice to be used as a metric to assess the improvement of the care delivery for sarcoidosis patients.

Using ICD codes alone to extract health information is far more convenient than the time-consuming manual review process of narrative datasets in unstructured data.[8] However, using ICD codes to identify sarcoidosis cases in large datasets with thousands of patients poses several practical challenges. First, given the heterogeneity of sarcoidosis, it is challenging to confirm the disease’s presence efficiently. The verification process requires careful analysis of the available narrative data such as progress notes, imaging reports, and pathology reports to establish the case definition based on the sarcoidosis diagnostic criteria.[3] Second, the precise identification of the type of organ involvement through EMR is a complex process and requires a thorough review of unstructured data. Although there are sub-codes for ICD diagnostic codes that aim to capture various organ involvement, healthcare providers may or may not be familiar with those sub-codes and may or may not use them correctly. Moreover, there are no specific ICD codes for classifying some of the various organ involvement in sarcoidosis (such as central nervous system or GI tract).[20] Third, ICD codes do not determine the extent of disease, such as that described by chest x-ray stages [21], due to a lack of ICD codes for different stages of pulmonary sarcoidosis.[20] Analysis of pulmonary features requires a manual review of every patient’s radiology reports and cannot be performed using only ICD codes alone. Finally, ICD codes do not specify the various sarcoidosis presentations such as acute, remitting, or chronic disease.[11,20] Thus, they cannot be used to classify patients into previously described phenotype groups.

The definition of clinical phenotypes has become an essential goal for the sarcoidosis scientific community, as genetic studies have identified different patterns of gene expression associated with disease severity and disease course.[22,23] In 2015, the National Heart, Lung, and Blood Institute (NHLBI) held a workshop to leverage current scientific knowledge and defining platforms to address disease disparities, identify high-risk phenotypes, and improve sarcoidosis outcomes [24]. Nine different steps and research strategies were recommended to expand the scope of sarcoidosis research, including EMR-based research to provide a unified, multidisciplinary approach to bring together stakeholders interested in reducing the burden and severity of sarcoidosis. However, the major barrier in the efficient use of EMR data is the accurate extraction of research-quality variables, case definitions, and outcomes.[25] Thus, the rapid identification of cases and extraction of relevant clinical variables from EMR using computable phenotype algorithms have emerged as an important next step in EMR-based research. Computable phenotype definitions are also essential to conducting pragmatic clinical trials and comparative effectiveness research, increasing the healthcare system’s capacity to deliver Precision Medicine effectively.[26]

The two most commonly applied approaches to defining computable phenotypes are: (1) a “*high-throughput”* phenotype algorithm using only structured data (traditionally, the ICD diagnosis codes). (2) a *“low-throughput”* phenotype algorithm that accesses structured and unstructured data to develop a sequential flow chart that should end with a case definition. Such a low-throughput approach employs high-performance computational tools (such as natural language processing [**NLP**]) to process text and extract information utilizing linguistic rules instead of labor-intensive manual review by researchers.[6] This approach should streamline the development of registries and help enrich EMR-based research studies.[27] Our study highlights the need for the development of such automated methods to improve the computational case-definition of sarcoidosis and other high-quality sarcoidosis-related research variables, such as determining the date of the diagnosis, organ involvements, Scadding stages, and the clinical status (acute, chronic, acute on the chronic or remitting disease).

Our study has several limitations. First, we used the ICD diagnosis code’s entered date as the surrogate time point to establish the patients’ care with active sarcoidosis or re-establish the care for those with remitting disease. This approach is different from what sarcoidosis researchers traditionally use as a surrogate to determine the date of diagnosis, which is the date of performing the biopsy. However, because the ICD diagnosis code’s date is more relevant for assessing the managing healthcare providers’ compliance, we chose to take the above approach. Second, in the cases where the biopsy report was unavailable (either due to a remote history of biopsy or biopsy performed outside the VA), we relied on the “secondary” histopathological reports documented in the providers’ narrative within the clinical notes. This approach made the diagnosis of sarcoidosis less robust because confirmatory biopsy reports in those patients were not directly verifiable. However, we used the index of suspicion approach to define probable sarcoidosis cases regardless of whether there was a confirmatory biopsy report available, which is consistent with the diagnostic algorithm recommended by the ATS practice guideline.[3] Last, the generalizability of our findings in VA EMR to other populations could be limited because the Veterans form a special population with different demographics and exposures from the general population. However, the US Veterans Affairs Healthcare System EMR data cover over 22 million Veterans across the US and over 14,000 patients with sarcoidosis ICD diagnosis codes, providing an enormous number of patients to study a rare disease.

## Conclusion

Although ICD codes can be used as reasonable classifiers to identify sarcoidosis cases within EMR, using computational algorithms to extract clinical and radiographic information (“index of suspicion”) from unstructured data could significantly improve case identification accuracy. Using automated emerging methods (such as NLP) to develop a novel sarcoidosis-specific computational phenotype algorithm could increase the efficiency of identifying these cases from large healthcare databases. Furthermore, we found that specialists are more likely than primary care providers to be “Fully” or “Substantially” compliant with the ATS clinical practice guideline. This finding calls attention to the importance of examining the association between physicians’ compliance with the recommendations and clinical outcomes longitudinally.

## Data Availability

Due to the sensitive nature of health data analyzed in the current study, data will remain confidential and are not publicly available.

## Ethics approval and consent to participate

The University of California San Francisco Institutional Review Board and the Veterans Health Administration Research and Development Committee approved this study. [IRB Protocol #15-16660].

## Acknowledgment

The Author(s) confirmed no figures or tables included from another publication.

